# Brief Report: Correlation cannot be used to compare sequencing panels used for the assessment of tumor mutational burden in non-small cell lung cancer

**DOI:** 10.1101/2020.03.28.20046102

**Authors:** Simon Heeke, Jonathan Benzaquen, Véronique Hofman, Elodie Long-Mira, Virginie Lespinet, Olivier Bordone, Charles-Hugo Marquette, Hervé Delingette, Marius Ilié, Paul Hofman

## Abstract

**Introduction:** Tumor mutational burden (TMB) has been proposed as a novel predictive biomarker for the stratification of patients undergoing immune-checkpoint inhibitor (ICI) treatment in non-small cell lung cancer (NSCLC) patients. The assessment of TMB has recently been established using large targeted sequencing panels and numerous studies are ongoing to harmonize the TMB assessment. However, usually “correlation” has been used to evaluate the association between the respective panels and we hypothesized that correlation might overestimate the comparability especially in lower TMB values, thus limiting the joint analysis of targeted sequencing panels for the assessment of TMB.

**Methods:** Thirty NSCLC samples from patients undergoing ICI treatment were consecutively sequenced using three large targeted sequencing panels: FoundationOne, Oncomine TML and QiaSeq TMB, respectively. TMB values were compared in the whole patient population and in a subset of patients where the TMB assessed by FoundationOne was between 5-25 mutations/Mb. Prediction of durable clinical benefit (DCB; >6 months no progression) was assessed using receiver operator characteristics and optimal cut-off values were calculated using Youden’s J.

**Results:** Correlation between the targeted sequencing panels was strong in the whole patient population between the three panels (R^2^ > 0.79) but was dramatically reduced in the subset of patients with TMB 5 – 25 mutations/Mb. All panels were able to predict DCB in the TMB high population.

**Conclusions:** Assessment of TMB using the three targeted sequencing panels was possible and predictive of response to ICI treatment but “correlation” was an inappropriate measurement to assess the association between the respective panels.

## Introduction

Tumor mutational burden (TMB) has been proposed as a novel biomarker for the prediction of response to immune-checkpoint inhibitors (ICIs) in non-small cell lung cancer (NSCLC) patients as well as in other cancer entities ^1,2^. While TMB has been initially assessed using whole exome sequencing (WES), targeted sequencing panels have been designed for the precise calculation of TMB as this is a more feasible approach for routine clinical practice ^3^. However, the assessment of TMB is not standardized across these panels, thus limiting the implementation of TMB in daily practice ^4^. We and others have compared the TMB between two targeted sequencing panels relying mainly on “correlation” as a mathematical measurement to determine the comparability of these panels and we could demonstrate good correlation between them ^4–6^. However, “correlation” is dominated by very high TMB values and consequently “accuracy” defined by the number of samples that are correctly determined to be TMB high has been proposed as a better measurement for the comparison of panels for TMB assessment ^7^. It is noteworthy that this proposition has been made using *in silico* analysis and has not been validated on real sequencing data ^7^. Here, we report for the first time the comparison of three different targeted sequencing panels used for the assessment of TMB in a real-life cohort of 30 NSCLC patients with a special emphasis on the precise selection of TMB-high patients.

## Materials & Methods

30 patients with advanced or metastatic lung adenocarcinoma who were treated with checkpoint inhibitors were included consecutively from routine clinical care (**Supplementary Table 1**). TMB was assessed using the Oncomine TML panel (OTML; Thermo Fisher Scientific, Waltham, USA) and the FoundationOne test (FO; Foundation Medicine, Cambridge, USA) as described previously ^5^. Additionally, the QiaSeq TMB panel (Qiagen, Hilden, Germany) has been used with the same DNA as used for the Oncomine TML panel, strictly following the manufacturer’s instructions and 40 ng of DNA was used for the sequencing runs. Prior to library preparation, DNA was treated with Uracil DNA glycosylase (UDG) to reduce artefacts introduced by deamination as published previously ^5^. Sequencing was performed on an Ion S5 sequencer (Thermo Fisher Scientific) and two samples were multiplexed per Ion 540 chip. Data was analyzed using CLC Genomics Workbench 12 (Qiagen). The different sequencing panels including number of genes are further highlighted in **Supplementary Table 2**. The number of identical genes used across the respective panels is shown in **Supplementary Figure 1**. Additionally, clinical data was collected, and clinical response was assessed using RECIST v1.1 criteria. Durable clinical benefit (DCB) defined by >6 months with no progressive disease has been assessed to define the cut-off values for the TMB-high population using receiver operator characteristics and Youden’s J. The study was performed in accordance to the guideline of the declaration of Helsinki and was approved by the local ethics committee (CHUN, IE-2017-905). All patients provided written informed consent.

## Results & Discussion

We analyzed the correlation between the two in-house tests (OTML and QiaSeq) to the outsourced FoundationOne assay (FO). As seen in **Figure 1A**, both in-house tests were well correlated to the FO assay with *R*^*2*^ = 0.819 for the OTML and *R*^*2*^ = 0.785 for the QiaSeq panel. However, we hypothesized that the correlation in the lower TMB range was much more relevant as the separation of TMB-high and TMB-low patients usually takes places at TMB values at around 10-15 mutations/Mb ^2,4,5,8^. We have previously established a cut-off of 15 Mutations/MB using the FO panel to determine the TMB high population ^5^. Consequently, we have analyzed the correlation of the panels only selecting samples where the TMB as determined by FO was between 5 and 25 mutations/Mb (± 10 Mutations/MB away from the cut-off) (**Figure 1B**). Interestingly, the *R*^*2*^ dropped dramatically to 0.0966 for OTML and 0.2453 for the QiaSeq panel, respectively (**Figure 1B**), demonstrating the very low correlation in this subset of tumors. It is noteworthy that for the in-house panels, a separate DNA extraction was used, compared to the FO panel where we had to send out tissue sections for the analysis. We tried to reduce the variation by using adjacent sections for the in-house extraction and FO but intra-tumoral heterogeneity might certainly explain some of the observed effects ^5,9,10^. Additionally, while the two in-house panels only use non-synonymous mutations for the calculation of TMB, the FO assay also includes synonymous mutations ^3^.

**Figure 1:**
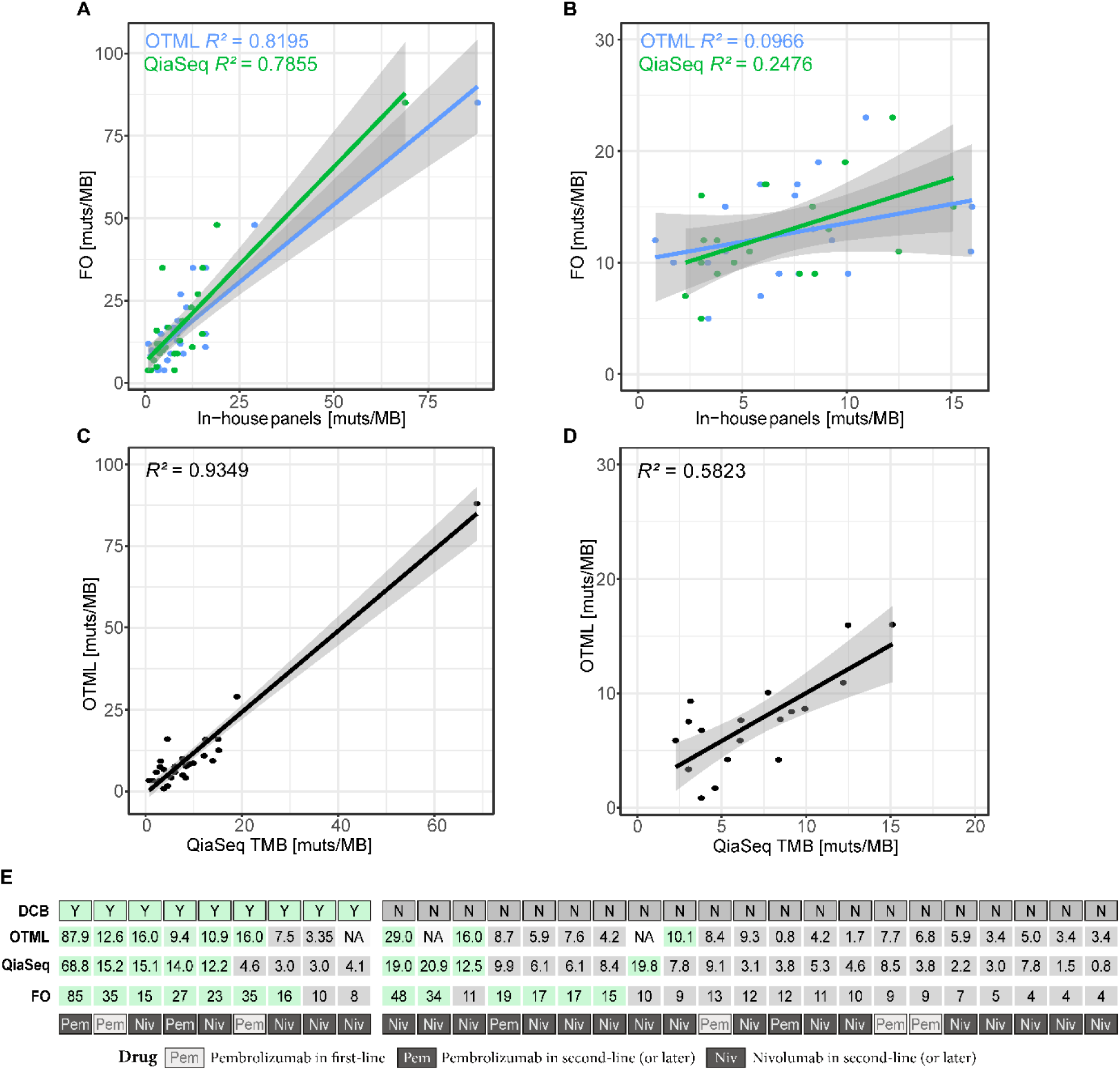
Correlation of Tumor Mutational Burden (TMB) across three targeted sequencing panels. **A)** The TMB of two in-house panels (Oncomine TML = OTML and QiaSeq TMB = QiaSeq) is correlated to the TMB obtained by the outsourced FoundationOne (FO) assay and *R*^*2*^ is given. **B)** Correlation of two in-house panels to FoundationOne in a subset of patients where the TMB as assessed by FO was between 5 and 25 mutations/Megabase (muts/Mb). **C)** Correlation of the two in-house tests against each other. **D)** Correlation of the two in-house tests in a subset of patients where the TMB as assessed by FO was between 5 and 25 muts/Mb. **E)** The durable clinical benefit (DCB) for each patient is given together with the TMB value obtained by the used targeted sequencing panels. TMB values that were classified to be TMB-high are highlighted in green. Lastly the respective immune-checkpoint inhibitor treatment is given in the last row.

Consequently, we also assessed the correlation of the two in-house panels where the same DNA was used for the sequencing. However, while the two panels seemed to be strongly correlated (**Figure 1C**), we also saw a dramatic drop of the correlation when filtering for patients with a TMB from 5-25 mutations/Mb (from *R*^*2*^ = 0.9349 to *R*^*2*^ = 0.5823; **Figure 1D**). Correlation values including the 95% confidence interval are further summarized in **Supplementary Table 3**. Poor agreement between pairs of panels using the Bland Altman method ^11^ can also be observed on **Supplementary Figure 2**, for the whole as well as the filtered populations. Interestingly, transformation of data using z-scores has been recently proposed to increase comparability between sequencing panels in an *in silico* approach, however, the authors noted that very high populations (>900 patients) are needed to perform this transformation with adequate statistical power ^12^ which was not possible in this cohort.

As correlation was clearly an inappropriate measurement to compare the different panels, we tested if TMB using the respective panels was able to predict response under ICI treatment in this series of NSCLC patients independently. All three panels predicted durable clinical benefit in this cohort with an area under the curve in ROC curves (AUROC) of FO = 0.8487, AUROC OTML = 0.7763 and AUROC QiaSeq = 0.6875 (**Figure 2A**). As expected, the cut-offs at which the TMB-high population was classified according to the Youden’s J analysis differed between the three panels with a cut-off of ≥ 14 mutations/Mb for FO, ≥ 9.3 mutations/Mb for OTML and ≥ 11.1 mutations/Mb for the QiaSeq panel, respectively (**Figure 2A**) ^6^. Likewise, progression-free survival (PFS) was prolonged in the TMB-high population (**Figure 2B**), independent of which panel was used. This indicates that, despite the low correlation between the panels, they were equally able to predict response to treatment, thus highlighting the fact that correlation cannot be used for the comparison of large sequencing panels used for TMB assessment. Consequently, only a subset of 6 patients was equally classified to be TMB-high across the three panels while several patients were classified as TMB-high in only a subset of the sequencing panels used (**Figure 1E**).

**Figure 2:**
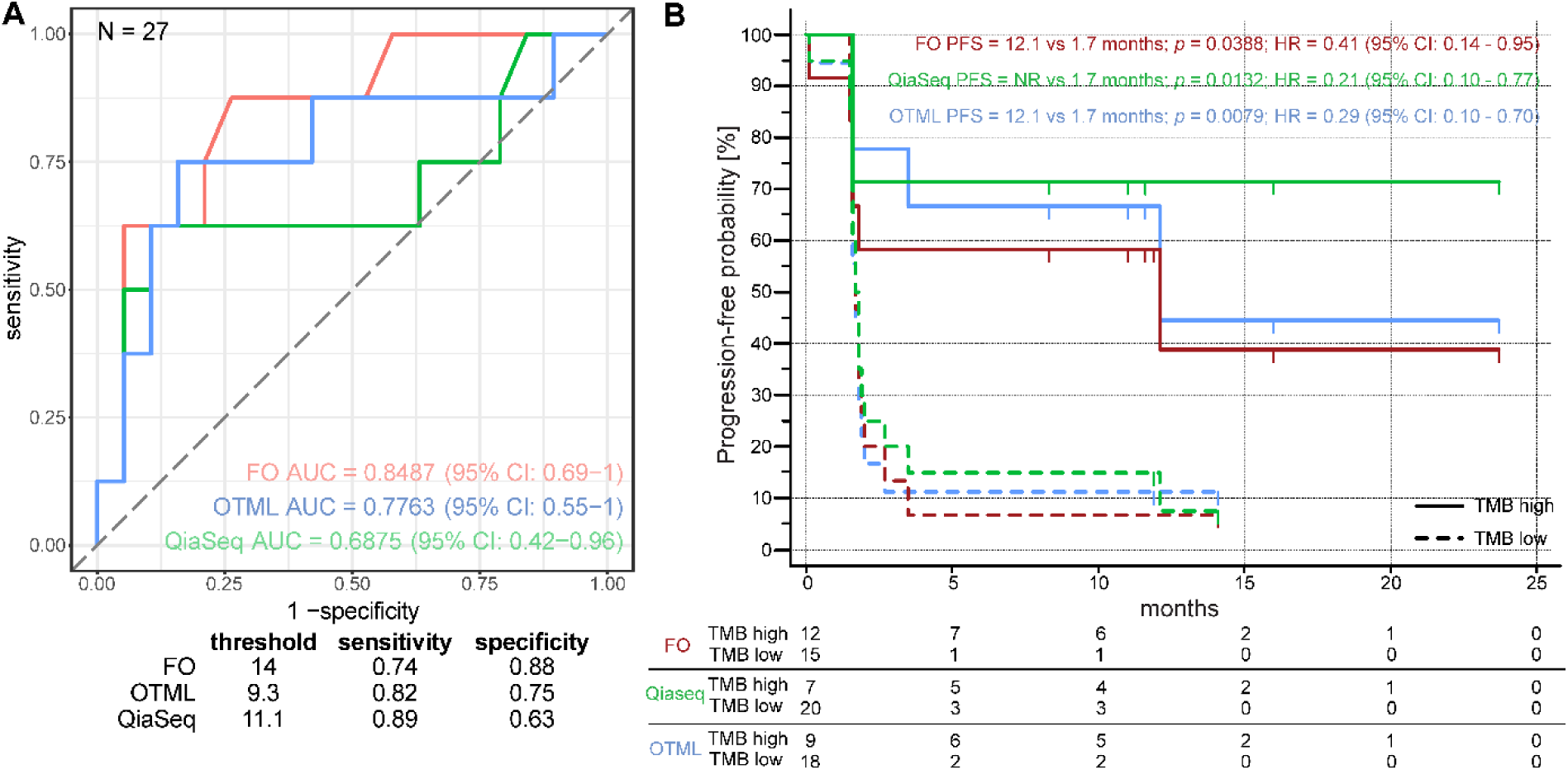
Clinical outcome of patients characterized by TMB. **A** Receiver Operator Characteristics (ROC) demonstrating the predictive performance of the targeted sequencing panels used. The area under the curve (AUC) together with the 95% confidence interval is given in the lower right corner. In total, 27 patients with successful sequencing data have been directly compared. The threshold was calculated using Youden’s J and is given in the table below together with sensitivity and specificity for each panel using the calculated cut-offs. **B** Kaplan-Meier plot for the assessment of progression-free survival. The TMB-high patients have been classified using the calculated cut-off in **A** and patients at risk are shown in the table below the figure. *p*-values were calculated using a log rank test and hazard ratio (HR) has been calculated using Cox Proportional Hazards Regression Analysis. 95% confidence interval (95% CI) is given for the Hazard ratio.

Taken together, these data demonstrate that the selection of patients undergoing ICI treatment based on TMB is dependent on the sequencing panel used, even though all panels are equally able to predict DCB in the cohort of patients. However, our study is limited by the inclusion of only a limited dataset of 30 patients and confirmation in larger and independent cohorts is crucial. There are several harmonization efforts underway to make TMB values comparable across different sequencing panels ^13^ and correlation has most often been used to demonstrate the comparison of different sequencing panels. However, the present study clearly demonstrates that the comparison of TMB values across different sequencing panels is inappropriate and that it might be preferable to assess the predictive performance of the respective sequencing panels independently of each other. Additionally, it seems critical to not only show the correlation of TMB values from the whole population but also from samples that are close to the calculated cut-off value to avoid overestimation of correlation based on very high TMB values.

## Data Availability

Data will be made accessible upon justified request.

## Acknowledgments

We would like to thank Abby Cuttriss from the Office of International Scientific Visibility at Université Côte d’Azur for proof reading.

## Supplementary Material

Supplementary Table 1: Patient Characteristics; Supplementary Table 2: Comparison of different gene panels used; Supplementary Table 3: Correlation assessment of different gene panels used. Supplementary Figure 1: Venn diagram highlighting overlapping genes used across the three sequencing panels; Supplementary Figure 2: Bland-Altman analysis of pairs of TMB values for the whole and the filtered populations.

